# Model-based evaluation of an infant HPV prophylactic vaccination program in Nigeria

**DOI:** 10.64898/2026.09.30.26364434

**Authors:** Robyn M. Stuart, Umeh Ifeoma Blessing, Christopher Gill, Jamie A. Cohen

## Abstract

**Background:** Nigeria introduced single-dose HPV vaccination for girls aged 9–14 in 2023, reaching approximately 60% aggregate coverage by 2025. While an important achievement, this remains short of the 90% coverage called for under the WHO’s 90-70-90 cervical cancer elimination targets, and further scale-up depends on reaching out-of-school girls. In this study we used a model of HPV transmission in Nigeria to quantify the residual cervical cancer burden under continued adolescent delivery, assess how additional screening scale-up and equity assumptions modify it, and identify the effective coverage requirements for an infant HPV vaccination program to be non-inferior to the existing adolescent strategy.

**Methods:** We used HPVsim, an agent-based microsimulation calibrated to Nigerian sexual behavior, HPV prevalence, and cervical cancer incidence, to project new cervical cancer cases and age-standardized incidence over 2025–2100 under scenarios which vary vaccination delivery mode (status-quo adolescent, WHO 90-70-90 adolescent scale-up, and 60/75/90% infant delivery), screening coverage (opportunistic 15% vs. scaled 70% aggregate), and the degree to which primary school completion correlated with vaccination (odds ratio 1 vs. 5) and with screening (odds ratio 1 vs. 3). Infant delivery scenarios were run at hypothetical effective-vaccine-efficacy-at-exposure values of 50%, 70%, and 95% — scenario inputs bracketing the plausible waning range, not evidence-based estimates of infant vaccine performance (no HPV vaccine trial has enrolled anyone below age 10). Each scenario was run at 50 replicates (10 calibration draws × 5 seeds); reported outcomes are the median across replicates and uncertainty ranges are interquartile (25th–75th percentile).

**Results:** Continuation of Nigeria’s current adolescent program is projected to avert 1.82 million cervical cancer cases (IQR 1.65–1.90 million; 95% UI 1.39–2.02 million) over 2025–2100 (a 53% reduction relative to no vaccination, IQR 51–55%; 95% UI 46–58%), with age-standardized incidence falling from 19.7 to 7.8 per 100,000 by 2100. However, 82% of the near-term (2025–2075) burden is projected to fall on the pre-2015 birth cohort, i.e. women already beyond the vaccination target age, leaving approximately 945,000 residual cases (IQR 905K–1.00M; 95% UI 835K–1.14M) in this cohort. Scaling up screening to WHO targets was estimated to avert a further ∼387K cases (IQR 331–424K; 95% UI 269–515K), roughly 25% of the residual burden; results were about 6–8% better if the scale-up instead reached all women equitably regardless of education. Under a hypothetical scenario of infant vaccination at 90% coverage and 95% effective vaccine efficacy at exposure (matching adolescent-age performance), outcomes were comparable to full WHO adolescent scale-up (49% vs. 54% reduction in vaccine-targetable cohort cancers), but under a hypothetical 50% effective efficacy at Nigeria’s DTP3-anchored 62% coverage the strategy averted only a fraction of these cancers.

**Conclusions:** The near-term cervical cancer burden in Nigeria is dominated by cohorts vaccination cannot reach; scaling screening for them is the more consequential lever over the coming three decades and rests on established evidence. Findings on infant HPV delivery are exploratory and hypothesis-generating: they depend on three unverified empirical prerequisites – infant HPV vaccine immunogenicity in the first months of life, durability of protection across the ∼15–25-year interval to peak HPV exposure, and coverage achievable through Nigeria’s routine childhood immunization platform – and should not be read as a basis for any policy recommendation contingent on infant HPV delivery until those prerequisites are established.

## Introduction

Despite effective prophylactic vaccines that prevent the acquisition of human papillomavirus (HPV), the causal agent of invasive cervical cancer, global coverage remains low, with only 33% of girls having received the HPV vaccine in 2025 (1). This figure stands in stark contrast to reported levels of vaccination coverage for infants and toddlers, with an estimated 84% of infants having received three doses of the vaccine against diphtheria, tetanus, and pertussis (DTP3) in 2023. One explanation for this significant discrepancy is the age of vaccination, with the HPV vaccine targeted to pre-adolescent girls, an age group that has not been traditionally reached with routine health care services. Many countries have leveraged school-based programs, and while such programs can be successful (2,3), they exclude out-of-school youth who also tend to be at higher risk for early sexual debut, concurrent sexually transmitted infections (STIs), and other conditions that might exacerbate their risk of HPV acquisition and persistence (4). Additionally, targeting girls just prior to sexual debut with a vaccine that prevents acquisition of an STI is potentially stigmatizing, and this has fueled vaccine hesitancy that limits uptake (5). In contrast, childhood vaccination series are a routine and established service that garner high levels of coverage even in the poorest and lowest resource settings.

There have been several studies to date examining the potential benefit of broadening the upper age of prophylactic HPV vaccination (i.e., catch-up vaccination for older girls and young women who were beyond the recommended age at the time of vaccine introduction) with evidence of decreasing marginal utility at older ages (6,7). The alternative direction, shifting the age of routine vaccination downwards into the childhood immunization series, is conceptually distinct and has received far less attention. For vaccines with limited duration of protection, vaccination should occur as close to exposure as possible, but a single dose of HPV vaccine has been shown to protect against HPV infection and, in one trial, against precancerous lesions, for up to roughly a decade in trials enrolling adolescents and young adults aged 15– 25 (8–12). While this evidence is not directly transferable to infants or to a cervical cancer endpoint, it is at minimum consistent with vaccination strategies targeted some years before exposure being biologically plausible.

Arguments in favor of an infant delivery approach include the theory that delivery costs may be lowered by adding to an existing platform, and that it may achieve higher/more equitable coverage than school-based delivery (delinking the vaccine from an STI vaccine could reduce the vaccine hesitancy associated with adolescent delivery, and using a delivery channel independent of school attendance would avoid the correlation between educational attainment and vaccine uptake that school-based delivery is prone to, and which may extend to the same women’s later engagement with screening). Arguments against this approach include the uncertainty regarding the ability to induce durable immune responses that will last until and beyond sexual debut and peak HPV exposure; cost of doses procured for infants on top of those for pre-adolescent girls; opportunity cost of not establishing and investing in a health care platform to reach pre-adolescent girls with health and social services they might not otherwise access; and the fact that routine childhood immunization platforms are not themselves guaranteed to reach children equitably.

Our study focuses on Nigeria, Africa’s most populous country with an estimated population exceeding 240 million and an estimated 12,000 new cases of cervical cancer and almost 8,000 cervical cancer deaths annually (13). Nigeria introduced HPV vaccination through Gavi support in October 2023 using a single-dose schedule of quadrivalent vaccine (HPV 16/18/6/11), targeting girls aged 9–14 through a phased rollout: an initial 15-state launch in October 2023 followed by expansion to the remaining 21 states plus the Federal Capital Territory in mid-2024 (14). School-based delivery has been the core modality, with community-based outreach through health facilities, door-to-door campaigns, and partnerships with community leaders reaching girls not currently enrolled in school. Complementary strategies are expensive to implement: the current program’s introduction and rollout costs are estimated at US$18.1 million, with a per-fully-immunized-girl cost of US$3.98 over a 5-year period (15).

Cervical cancer control in Nigeria is further constrained by the near-absence of population-based secondary prevention: Nigeria lacks a tailored national cervical cancer control policy or population-based screening program, and coverage of the opportunistic screening that does exist reaches only about 10% of women in their lifetime (16). Vaccination cannot retroactively protect the large share of Nigerian women who are already past the target age for HPV vaccination, so screening is the only lever available for this group; the burden of disease over the next several decades therefore depends on what can be achieved through both primary prevention (vaccination) for future cohorts and secondary prevention (screening) for cohorts vaccination cannot reach. The next critical stages for Nigeria are therefore: (i) scaling screening to reach the cohorts vaccination has already missed; (ii) maintaining or scaling the vaccination coverage achieved during the initial 9–14 rollout, especially among out-of-school girls who are otherwise excluded from school-based delivery; and (iii) considering alternative delivery strategies that could reach a broader fraction of future birth cohorts. It is worth investigating what level of efficacy would be required for an infant HPV vaccine to be competitive with the existing strategy of vaccinating 9–14 year olds, a strategy that has demonstrated high efficacy but faces difficulties in sustaining high coverage, especially among girls at greatest risk.

In this study we therefore examined three complementary levers for reducing Nigeria’s cervical cancer burden: (1) sustaining and scaling the current adolescent vaccination program; (2) scaling opportunistic screening, the only intervention available to birth cohorts vaccination has already missed, and one that may itself be limited by the same socioeconomic determinants that shape vaccination uptake; and (3) shifting vaccine delivery to infancy for future birth cohorts, which trades adolescent vaccination’s demonstrated efficacy and delivery track record for the potential benefits of a routine childhood delivery channel, evaluated here in terms of its likely uptake by age group and its efficacy over the long interval between infant vaccination and peak HPV exposure. Our analysis estimated total cervical cancer cases averted across a range of scenarios spanning these three levers.

## Methods

### Model overview and Nigeria calibration

We used HPVsim (v3.1.0), an open-source agent-based model of HPV transmission and cervical disease progression described in detail elsewhere (17), to build a simulated population that lets us run and compare vaccination and screening scenarios for Nigeria under identical conditions. Our HPVsim model for Nigeria starts by simulating a population with the demographics of Nigeria (18), in which HPV is transmitted through marital and casual partnerships. Four HPV genotype categories are modeled explicitly: HPV16 and HPV18 individually, plus two pooled high-risk oncogenic groups: Hi5 (the five additional high-risk types included in the 9-valent vaccine: 31, 33, 45, 52, 58) and other-HR (five high-risk types not covered by the 9-valent vaccine: 35, 39, 51, 56, 59). HPV infections progress through pre-cancerous (pre-CIN and CIN) states with genotype- and age-specific transition rates, and a small fraction of persistent infections progress to invasive cervical cancer. Nigeria-specific demographic dynamics are captured using standard age- and sex-stratified fertility and mortality data, so that projected population growth to 2100 is reflected in counterfactual denominators.

Because school attendance shapes how HPV vaccination currently reaches girls in Nigeria, we also modelled each girl’s schooling directly: girls start primary school around age 6, though many enter late, and some leave before finishing, so that by age 9 roughly 70% of girls are still in school and 30% are not, consistent with national household-survey estimates of primary attendance and out-of-school rates (19,20). Whether a girl completes primary school affects her chances of being vaccinated and later screened (see below).

We calibrated the model to Nigeria using the same methodology previously applied to India (17), Tunisia (21), and to 30 sub-Saharan African countries (22). Calibration was performed by using a Bayesian optimization algorithm to search over the 14 free parameters listed in Table S1 and minimize a goodness-of-fit score, defined as the weighted sum of squared differences between simulated and observed values across five target series: age-specific new cervical cancer cases (Globocan 2020), age-standardized cancer incidence (WHO 2000 standard), HPV genotype distribution in cancer, HPV genotype distribution in high-grade lesions, and age-specific HPV DNA prevalence in women aged 15–49 (HPV Information Centre meta-analyses). Series weights balance the number of data points per series so that no single high-density series dominates the objective.

The search produced 1000 candidate parameter sets. We ranked these by goodness-of-fit score and retained the top 50 as our calibration posterior sample (Table S1 reports the range across this sample; Figure S3 overlays their time series against the target data). To propagate calibration uncertainty into scenario runs, we sampled 10 draws from this posterior sample and ran each across every scenario at 5 different model seeds, producing 50 replicates per scenario (see *Uncertainty and reporting*, below). This captures calibration uncertainty (variation across the 10 draws) and stochastic simulation uncertainty (variation across the 5 seeds) jointly in the reported bands. Figure S2 is the primary goodness-of-fit diagnostic, showing the fit of the top-ranked parameter set against age-specific cancer counts, age-standardized incidence, and genotype-distribution targets.

### Cervical cancer prevention

Nigeria’s current approach to cervical cancer prevention rests on adolescent vaccination and opportunistic screening/treatment. We modeled each to reflect how it actually operates today, before varying them in our scenarios.

Nigeria delivers HPV vaccination as a single dose of quadrivalent vaccine (protecting against HPV16, 18, 6, and 11), supplied through Gavi, to girls aged 9–14, primarily through schools with catch-up outreach for girls not enrolled. We modeled this directly: the vaccine confers 95% efficacy against HPV16/18 at the time of vaccination under our base-case assumption, plus cross-protection against non-vaccine types taken from published estimates for the quadrivalent vaccine.

Nigeria has no dedicated national cervical cancer screening program; screening that does happen is opportunistic and reaches an estimated 10–15% of women in their lifetime (16). Where screening does occur, most women (∼80%) are screened with visual inspection using acetic acid (VIA), a low-cost test that allows same-day treatment (typically thermal ablation) for women found positive; the remainder are screened with HPV DNA testing, which is more sensitive but requires a separate follow-up visit (VIA or colposcopy) to confirm a positive result before treatment can start, a step that in practice only ∼20% of HPV-positive women complete (23).

Both vaccination and screening are shaped by education. Girls in school are substantially more likely to be vaccinated than girls who are not, so we set the vaccination odds ratio between in-school and out-of-school girls to 5, consistent with in-school vs. out-of-school HPV coverage gaps reported from comparable school-based campaigns in Malawi and Ethiopia (edu_OR ≈ 5–12) (24,25); we are not aware of any study that has directly measured this gap for Nigeria’s own HPV program, so we use the regional campaign evidence as the best available proxy. Screening uptake in Nigeria is also stratified by education. A population-based study of Nigerian women aged 30–49 (26) reports screening rates stratified by education level; pooling everyone who completed at least primary school against those with none gives an odds ratio of approximately 3. We use this as our target assumption for the education-linked screening gap.

### Effective coverage, equivalence, and treatment of waning

Whether infant vaccination compares favorably to adolescent vaccination depends on the effective coverage of each program at the time of HPV exposure, i.e. the product of vaccine coverage and vaccine efficacy at exposure. Under stable duration of protection, a shift from adolescent to infant delivery yields equivalent projected cervical cancer burden under the model whenever

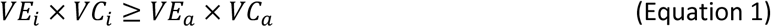

where subscripts *a* and *i* denote adolescents and infants respectively, VE denotes vaccine efficacy against acquisition at exposure, and VC denotes coverage of the target cohort. Figure 1A plots the coverage/efficacy isocurves implied by Equation 1 assuming an adolescent-delivery efficacy of 95% and varying adolescent coverage (VCA) between 10% and 90%.

**Figure 1:**
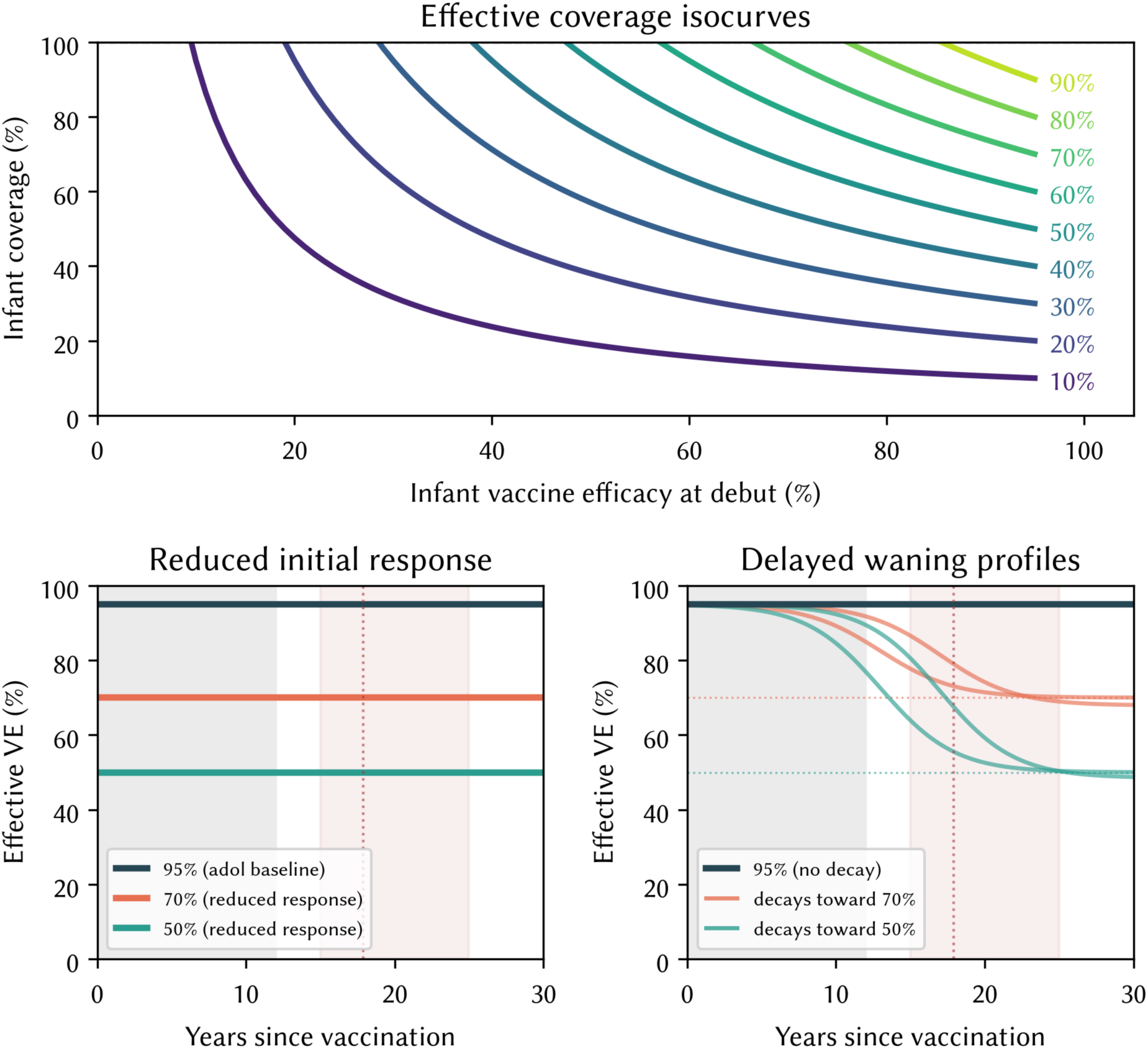
Analytical framing for infant HPV vaccine delivery. (A) Required infant vaccine coverage as a function of infant vaccine efficacy at debut, for adolescent-equivalent coverage targets of 10–90% (colored curves), assuming 95% adolescent efficacy (Equation 1); the dashed line marks 100% coverage, the maximum theoretically achievable. (B) Illustrative waning mechanism (a): an immature infant immune system produces a genuinely lower initial response (70% or 50%, orange/teal) than the adolescent benchmark (95%, navy), which does not itself decline over time. (C) Illustrative waning mechanism (b): the infant response initially matches the adolescent benchmark, holds flat through the ∼12-year window over which current immunogenicity studies have followed vaccinated cohorts (grey shading), then declines before Nigeria’s peak HPV exposure window (pink shading; dotted line marks the median age at first sex).

Because infant vaccination is administered in the first months of life while peak HPV exposure in Nigeria follows sexual debut at a median age of 17.9 years (27), with cumulative exposure risk continuing to build through the mid-20s, durability of protection across this ∼15–25-year gap is the central biological uncertainty for this delivery strategy. Rather than fit an explicit waning function inside HPVsim, we take advantage of the observation that any biological explanation for a durability shortfall reduces to a single quantity: the effective VE remaining at exposure, the fraction of administered efficacy that persists to the age each cohort acquires HPV. Two qualitatively distinct mechanisms could produce the same shortfall relative to the ∼95% adolescent-equivalent benchmark (Figure 1B, C): (a) an immature infant immune system could mount a genuinely lower initial response that does not itself wane; or (b) the infant response could match the adolescent benchmark and hold flat through the ∼12-year window over which currently available immunogenicity studies have followed vaccinated cohorts (8–12), then decline before the exposure window opens. Because the evidence window (≤12 years) ends only a few years before the exposure window begins, no existing study can distinguish between these mechanisms, or between either of them and no shortfall at all. In our simulations we do not choose between mechanisms; instead we treat the effective VE at exposure as the value we varied, testing 50%, 70%, and 95% for infant delivery, values chosen to bracket the plausible range implied by both mechanisms without requiring a specific parametric form. This framing lets us report our results as requirements on effective coverage, agnostic to mechanism.

### Scenario design

We designed the scenarios listed in Table 1 to answer three questions in turn: how much of Nigeria’s cervical cancer burden is likely to be addressed by continuation of current vaccination programs (addressed by comparing status quo to no vaccination); how much of the remaining burden screening alone could avert, and whether that depends on delivering screening equitably (addressed by the screening scale-up group, which holds vaccination fixed); and how an infant delivery strategy compares once vaccination itself is also scaled or shifted in delivery mode (addressed by the WHO scale-up and infant delivery groups). All scenarios share Nigeria’s historical baseline: an opportunistic screening program covering ∼15% of women aged 30–50 over their lifetime (16), and the 2023–2025 HPV vaccination rollout targeting girls aged 9–14 at aggregate coverages of 27%, 60%, and 60% respectively. Scenarios diverge from 2026 onward. Full specifications of each scenario including all parameters and assumptions are given in Table S2.

**Table 1.** Scenarios modeled.

| Scenario | Adolescent vaccination (2026+) | Infant vaccination (2030+) | Lifetime screening coverage in women 30-50 | Vaccination education gap | Screening education gap |
| --- | --- | --- | --- | --- | --- |
| No vaccination | None | None | 15% | — | — |
| Status quo | 60% | None | 15% | edu_OR = 5 | edu_OR = 3 |
| Status quo vaccination with screening scale-up (correlated with education) | 60% | None | 70% | edu_OR = 5 | edu_OR = 3 |
| Status quo vaccination with screening scale-up (equitable) | 60% | None | 70% | edu_OR = 5 | edu_OR = 1 |
| WHO targets | 90% | None | 70% | none | edu_OR = 3 |
| Infant delivery, 95% efficacy | 60% (2026-2029 bridge only) | 60%, 75%, 90% | 70% | none (uniform) | edu_OR = 3 |
| Infant delivery, 70% efficacy | 60% | 60%, 75%, 90% | 70% | none (uniform) | edu_OR = 3 |
| Infant delivery, 50% efficacy | 60% | 60%, 75%, 90% | 70% | none (uniform) | edu_OR = 3 |

All scenarios share Nigeria's 2023–2025 historical rollout before diverging in 2026 as shown above. Infant scenarios continue the status-quo adolescent program as a bridge through 2029, for birth cohorts too old for infant delivery but too young to have been reached by the routine adolescent program, before switching over to infant delivery in 2030.

To answer the first question, we compare the status-quo continuation scenario (adolescent vaccination continues at 60% aggregate coverage post-2026 with primary school completers 5x more likely to be vaccinated; screening remains at 15% coverage) with a no vaccination counterfactual.

To address our second question, we isolate the effect of screening by holding vaccination fixed at the status-quo 60% aggregate coverage and varying how screening scales beyond 2026. In addition to the status-quo scenario, we model a scale-up scenario reaching the WHO 90-70-90 elimination target of 70% lifetime screening coverage in women aged 30–50 [WHO, 2020], again with an odds ratio of 3 differentiating screening uptake among primary-completers vs non-completers; the underpinning modelling analysis (Brisson et al., 2020) applied this same 70% figure in a 78-country analysis that included Nigeria. We also include a scenario with the same screening scale-up but with an odds ratio of 1 to estimate how much of the outcome depends on the education gap.

Finally, to explore the impact of fully-scaled vaccination delivery, we run a scenario in which adolescent vaccination scales to the WHO target of 90% coverage in both in-school and out-of-school girls (i.e. we assume no vaccination-education gap for this scenario) while screening scales to 70%. We contrast this with a scenario in which infant vaccination replaces adolescent vaccination, with a single-dose infant program from 2030 onwards at 90% coverage uniformly across the birth cohort, and screening scaled up to 70%. These two scenarios are designed so that the only difference between the adolescent and infant delivery strategies is in the target age, with both strategies attaining high and equitable vaccination coverage plus high (albeit education-stratified) screening coverage. This comparison is intended to show the circumstances under which the infant delivery strategy would be directly equivalent to existing strategies. In reality, however, there are many unknowns about how infant delivery would function. We therefore also run additional infant delivery scenarios at 3 effective-VE-at-exposure values (95%, 70% 50%), spanning the plausible waning bounds described in Figure 1, and 3 coverage levels (60%, 75%, and 90%).

### Education-uptake sensitivity analysis

The vaccination and screening education odds ratios in Table 1 (vax_OR = 5 and screening_OR = 3 respectively) are single-point assumptions drawn from a mix of regional (vaccination) and Nigeria-specific (screening) evidence, and are important drivers of our equity conclusions. Rather than treating them as fixed nuisance parameters, we built the sensitivity to these ORs into the scenario matrix itself: for each scale-up scenario in Table 1, we run a version with the assumed OR and a paired version with OR = 1 (no education gap). The screening_OR contrast (3 vs. 1) is isolated by the two “screening scale-up” rows of Table 1, holding vaccination at status-quo 60% aggregate with vax_OR = 5. The vax_OR contrast (5 vs. 1) is bundled inside the WHO scale-up row, which raises aggregate coverage to 90% and simultaneously closes the education gap — the two are deliberately coupled because reaching 90% aggregate coverage in Nigeria requires closing the in-school/out-of-school gap, so treating them as separable would be misleading. The infant delivery scenarios similarly assume uniform coverage across the birth cohort (i.e. vax_OR = 1), which functions as a further contrast against the status-quo vax_OR = 5 baseline.

To probe intermediate OR values, we also ran a finer sweep: vaccination OR ∈ {2, 3, 8} at status-quo 60% aggregate coverage with baseline screening, and screening OR ∈ {2, 5} at the 70% scale-up target with status-quo vaccination. Together with the paired-OR contrasts above, this gives full sweeps across vax_OR ∈ {1, 2, 3, 5, 8} and screening_OR ∈ {1, 2, 3, 5}. Results are reported in Supplementary Figure S4 (cancers averted vs. OR value, one panel per gradient) and Supplementary Table S3 (numeric results per OR value).

### Uncertainty and reporting

To quantify uncertainty, we ran each scenario at every combination of 10 parameter sets and 5 seeds, giving 50 replicates per scenario. Parameter sets were drawn from the top of the calibration posterior (ranked by goodness-of-fit score, as described above). Reported outcomes are the median across replicates; uncertainty ranges shown in figures and quoted in the text are the interquartile range (IQR, 25th–75th percentile) and, for headline results, the 95% uncertainty interval (UI, 2.5th–97.5th percentile) across the 50 replicates. These intervals reflect the joint spread across the 10 calibration draws and the 5 seeds within each draw; they are empirical spread across replicates, not posterior credible intervals in a Bayesian sense. For each simulation, we recorded new cervical cancer cases by year and by birth cohort, tracking women born before 2015 separately from the six 5-year birth cohorts spanning 2015–2044, plus HPV prevalence, ASR of cancer incidence, and cases stratified by vaccination × screening status at the time of diagnosis, over the period 2025–2100. Cumulative outcomes are computed to 2100 for the primary results and to 2125 for the pre-2015 cohort tail, which extends beyond 2100 for women alive in 2020.

## Results

Under a continuation of the current adolescent vaccination program at 2025 coverage levels (approximately 60% aggregate coverage among girls aged 9–14, with an education-linked coverage gap), and with existing opportunistic screening reaching roughly 15% of women aged 30–50 in their lifetime, we estimate that 1.82 million cervical cancer cases (IQR 1.65–1.90 million; 95% UI 1.39–2.02 million) would be averted over 2025–2100 relative to a no-vaccination counterfactual (Figure 2A), a reduction of 53% (IQR 51–55%; 95% UI 46–58%). We estimate that age-standardized cervical cancer incidence would fall from 19.7 per 100,000 in 2025 to 7.8 per 100,000 by 2100 (IQR 6.1–9.0; 95% UI 2.2–10.7) vs. 25.8 (IQR 23.7–28.7; 95% UI 16.8–31.0) under no vaccination, consistent with the entry of vaccinated cohorts into their peak-risk age band from the mid-2040s onward.

**Figure 2:**
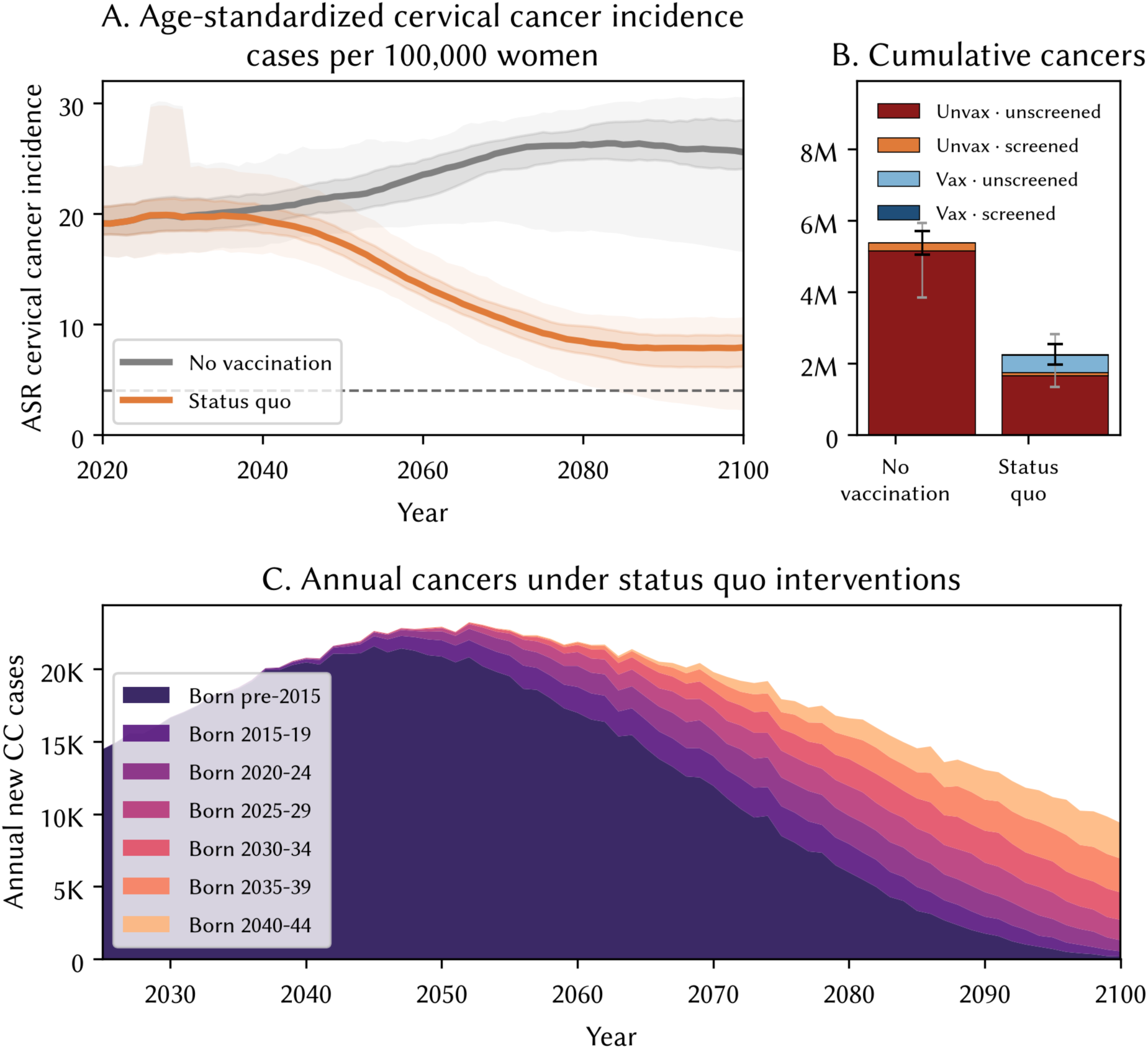
Status-quo cervical cancer burden under continued adolescent vaccination. (A) Age-standardized cervical cancer incidence (cases per 100,000 women) 2020–2100 under no vaccination (grey) versus status-quo continuation of the current adolescent program (orange); inner shaded band = interquartile range (25th–75th percentile), outer band = 95% uncertainty interval (2.5th–97.5th percentile) across the 50 replicates (10 calibration draws × 5 seeds); dashed line marks the WHO cervical cancer elimination threshold (4 per 100,000). (B) Cumulative cervical cancer cases 2020–2125, stratified by vaccination and screening status at diagnosis; whiskers show IQR (thick, black) and 95% UI (thin, grey) on the total. (C) Annual new cervical cancer cases under status-quo interventions, decomposed by birth cohort.

However, the near-term cervical cancer burden in Nigeria is dominated by women who were already beyond the target age for vaccination when the program launched in 2023. 82% of the cervical cancers projected between 2025 and 2075 (IQR 80–83%; 95% UI 78–87%) occur in women born before 2015 (which we hereafter refer to as the pre-2015 cohort) with the remainder distributed across the six vaccine-targetable birth cohorts (born 2015–2044). Vaccinating girls aged 9–14 has only a small direct effect on this pre-2015 group (Figure 2B, C): approximately 945,000 cancers (IQR 905K–1.00M; 95% UI 835K– 1.14M) over 2025–2100 arise among women born before 2015 even under continued vaccination (down from 1.14 million with no vaccination, IQR 1.09–1.19M; 95% UI 1.01–1.30M — a 16% reduction, IQR 15– 17%; 95% UI 11–20%, from indirect and cohort-edge effects). Reducing this residual burden depends primarily on the reach of screening and treatment among these women during the remainder of their time in the 30–50 screening window.

For the six vaccine-targetable birth cohorts (born 2015–2044), continued adolescent vaccination at status-quo coverage would reduce total lifetime cervical cancer cases from approximately 1.72 million (IQR 1.63– 1.79 million; 95% UI 1.26–1.87 million, no vaccination) to 549,000 (IQR 466–631K; 95% UI 258–708K), an aggregate reduction of 68% (IQR 65–72%; 95% UI 60–79%) across the six cohorts, ranging from 63% (IQR 58–67%; 95% UI 53–72%) in the 2015–2019 cohort to 70% (IQR 65–74%; 95% UI 61–86%) in the 2040–2044 cohort (which experiences the full mature program from age 9). Figure 2C shows the annual cases from each cohort year by year: the six vax-targetable cohorts remain a relatively small share of annual burden through 2075, then dominate as the pre-2015 cohort ages out.

Scaling opportunistic screening to a WHO-consistent lifetime coverage target of ∼70% among women aged 30–50, with the education-correlated gap we take as our core assumption (odds ratio 3), averts an additional ∼387,000 cancers (IQR 331–424K; 95% UI 269–515K) over 2020–2125 (∼25% of the residual burden after scaling up vaccination to 60% coverage, IQR 24–26%; 95% UI 21–28%), of which 216K (IQR 204–239K; 95% UI 169–289K) are in the pre-2015 cohort and 163K (IQR 127–189K; 95% UI 66–251K) are in the vaccine-targetable cohorts (Figure 3B). Even so, ∼805,000 pre-2015 cancers (IQR 756–835K; 95% UI 713–949K) remain even under the fully scaled screening program; the residual reflects the fraction of women who age out of the 30–50 screening window before 2026, plus women reached only late in that window, plus test/treatment cascade losses.

**Figure 3:**
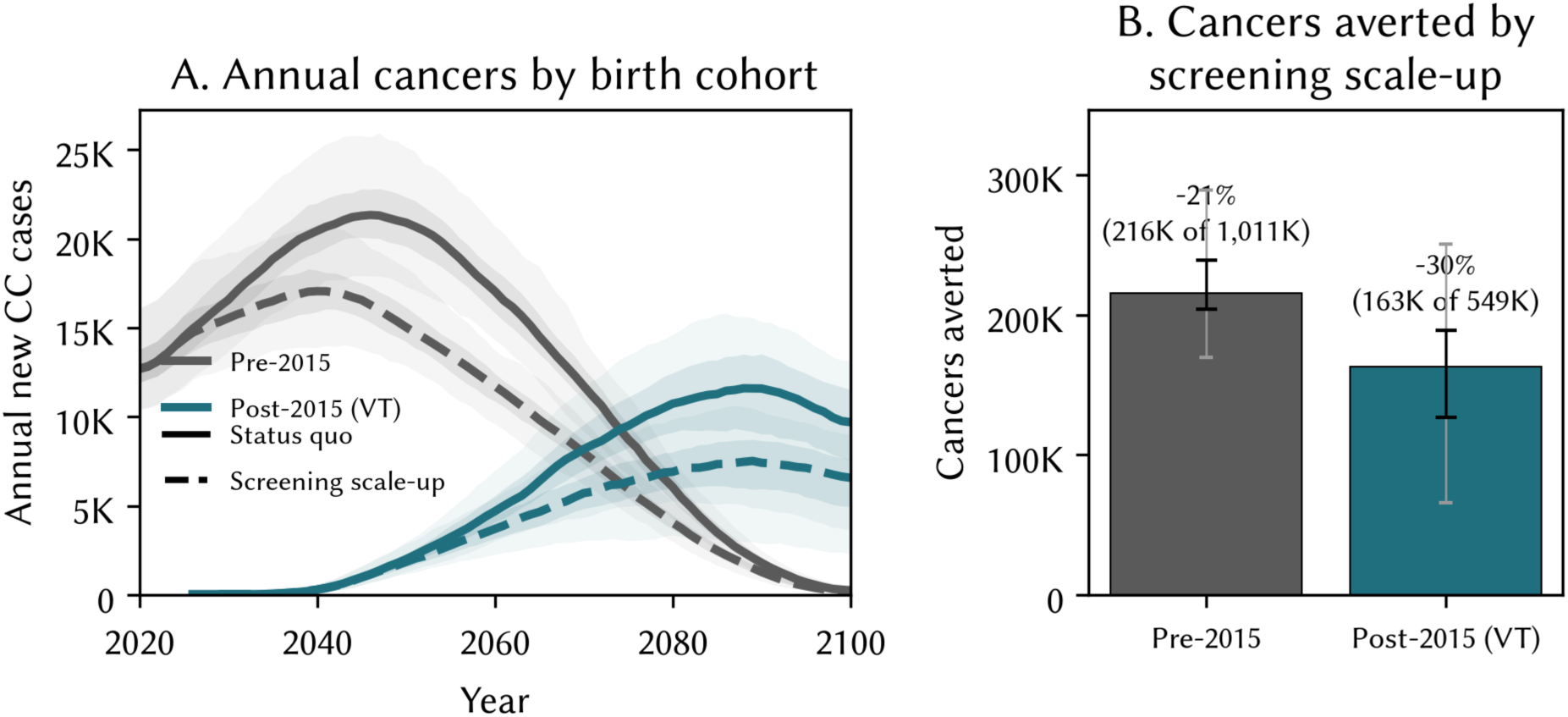
Screening scale-up redistributes averted cancers by birth cohort. (A) Annual new cervical cancer cases 2020–2100 for the pre-2015 cohort (grey, ineligible for adolescent vaccination) and the pooled post-2015 vaccine-targetable cohorts (teal), each under status quo (solid) and under screening scaled up to a 70% lifetime coverage target with the education-correlated gap we take as our core assumption (dashed); inner shaded band = IQR (25th–75th percentile), outer band = 95% UI (2.5th–97.5th percentile) across the 50 replicates. (B) Cumulative cancers averted by screening scale-up over 2020–2125 for each cohort group, with the percentage and absolute number averted relative to the status-quo total; whiskers show IQR (thick, black) and 95% UI (thin, grey).

This result assumes screening scale-up reproduces the same education-correlated gap we assume for status-quo vaccination. If instead the scaled-up program reached primary-completers and non-completers equitably at the same 70% aggregate coverage, outcomes are only modestly better: about 8% fewer cervical cancers in the vaccine-targetable cohorts (∼30K fewer cases, IQR 13–51K; 95% UI −5 to 76K) and about 6% fewer in the pre-2015 cohort (IQR 5–7%; 95% UI 2–9%).

The full education-uptake sensitivity across finer OR values is shown in Supplementary Figure S4. For the vaccination gradient at status-quo 60% aggregate coverage, cancers in vaccine-targetable cohorts are essentially insensitive to the vaccination OR across the range {2, 3, 5, 8} (all differences within noise; Fig S4A), because holding aggregate coverage constant only redistributes vaccination between in-school and out-of-school girls without changing the total number vaccinated — the OR shapes the *distribution* of residual burden across education groups, not the aggregate. For the screening gradient at the 70% scale-up target, cancers averted in vaccine-targetable cohorts decline monotonically with the screening OR: from ∼153K (IQR 121–190K; 95% UI 71–222K) at OR = 1 (fully equitable) to ∼117K (IQR 87–139K; 95% UI 54–173K) at OR = 5 (a larger gap than we assume), with the OR = 3 baseline at ∼127K (Fig S4B).

Fully scaling both vaccination and screening to WHO 90-70-90 targets, with the education-correlated screening gap as our core assumption, would avert ∼472,000 cancers (IQR 433–525K; 95% UI 341–583K) over 2025–2100 relative to status quo, including a 54% reduction (IQR 52–57%; 95% UI 47–64%) in cancers among the vax-targetable birth cohorts (Figure 4). However, reaching this scenario requires vaccinating 90% of adolescent girls, which in turn requires reaching out-of-school girls at the same coverage as those in school, an assumption that is not currently met and that, if unrealized, would leave overall adolescent coverage at approximately 60% (i.e. a continuation of status quo).

**Figure 4:**
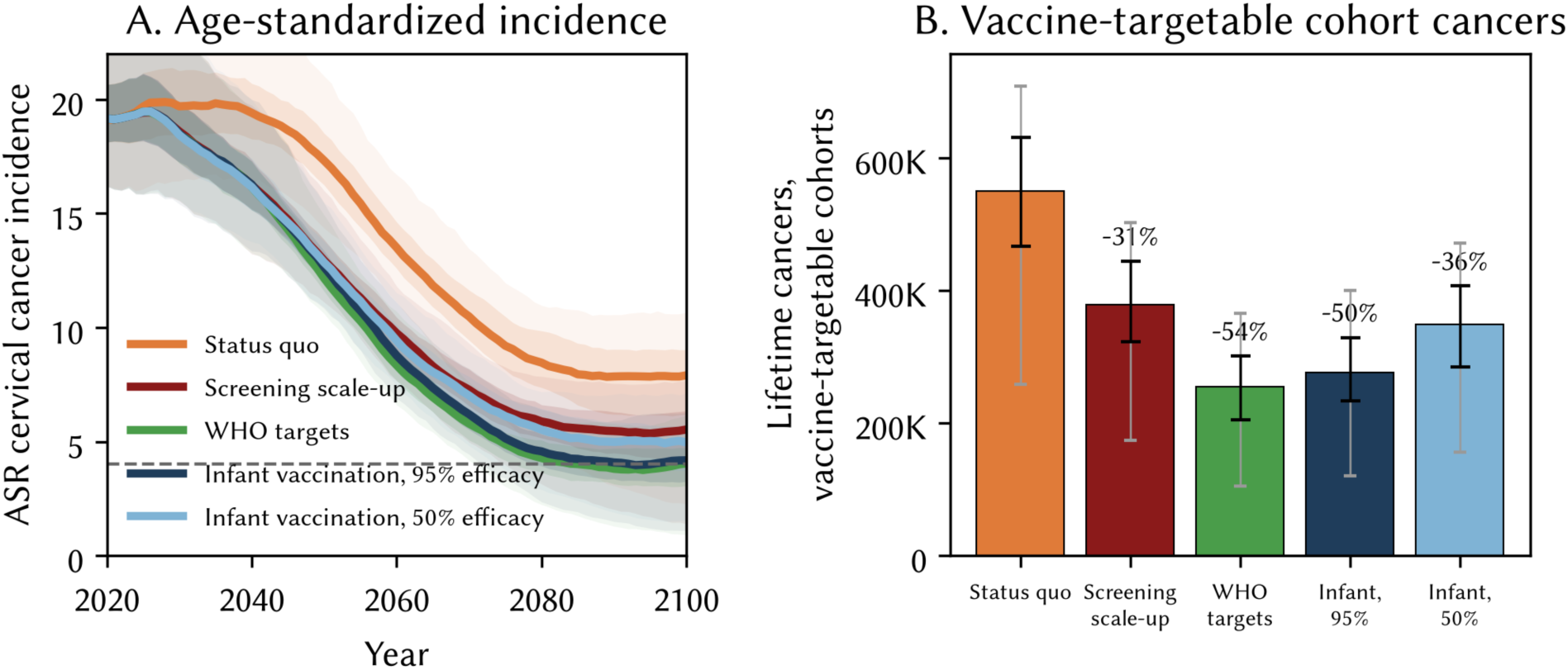
Infant delivery compared with screening and adolescent-vaccination scale-up. (A) Age-standardized cervical cancer incidence 2020–2100 under status quo, screening scale-up alone, full WHO 90-70-90 scale-up (adolescent vaccination and screening both raised to target), and infant delivery at hypothetical 95% and 50% effective vaccine efficacy at exposure (scenario inputs bracketing the plausible waning range, not evidence-based estimates); inner shaded band = IQR, outer band = 95% UI across the 50 replicates; y-axis truncated at 22 per 100,000 for readability (some outer bands extend slightly above this). Dashed line marks the WHO cervical cancer elimination threshold (4/100,000). (B) Lifetime cervical cancer cases 2020–2125 in the vaccine-targetable birth cohorts under each scenario, with the percentage reduction relative to status quo; whiskers show IQR (thick, black) and 95% UI (thin, grey).

Infant delivery offers an alternative route to high coverage that does not depend on school attendance. Under a hypothetical baseline scenario in which infant vaccine efficacy matches adolescent efficacy (95% at exposure — a scenario input, not an evidence-based estimate), a program reaching 90% of infants achieves outcomes broadly comparable to full WHO scale-up: 219,000 cancers (IQR 182–253K; 95% UI 100–315K) in the vax-targetable cohorts vs. 194,000 (IQR 158–227K; 95% UI 87–283K) under the WHO scale-up scenario. The remaining gap reflects the fact that the WHO scale-up scenario catches older adolescent cohorts (born 2009–2016) that fall between the historical adolescent program and the 2030 infant switch-over.

The strength of the infant strategy erodes if either coverage falls below 90% or efficacy falls below the adolescent benchmark (Figure 5). Under the sensitivity grid’s low corner — 60% coverage (comparable to Nigeria’s DTP3 coverage levels) and hypothetical 50% effective VE at exposure (a scenario input representing either a less effective vaccine or rapid waning between infant vaccination and peak HPV exposure) — infant delivery averts only 15% of lifetime cancers in vax-targetable cohorts. Combining Equation 1 with the observed adolescent program efficacy (∼95%) suggests that an infant program would need effective VE at exposure ≥ 63% at 90% coverage, or ≥ 90% at 60% coverage, to be non-inferior to Nigeria’s current 60% aggregate adolescent coverage.

**Figure 5:**
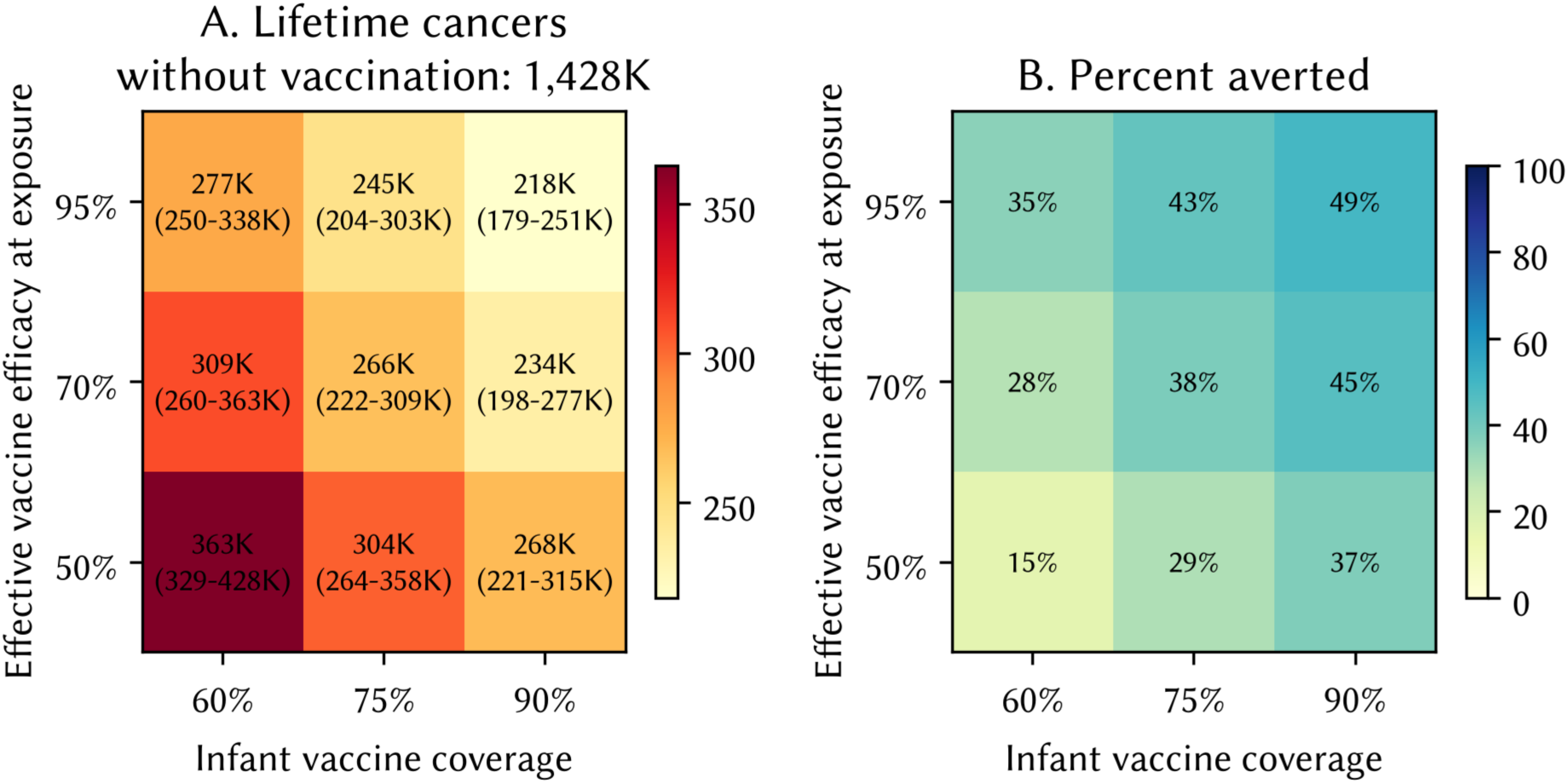
Infant delivery outcomes across the coverage-by-efficacy sensitivity grid. (A) Lifetime cervical cancer cases in the vaccine-targetable birth cohorts (2025–2100) for infant vaccination at coverage levels of 60%, 75%, and 90% (columns) and hypothetical effective vaccine efficacy at exposure of 50%, 70%, and 95% (rows) — scenario inputs bracketing the plausible waning range, not evidence-based estimates. (B) The same grid expressed as the percentage of cases averted relative to status quo.

## Discussion

Using a Nigeria-calibrated HPV transmission model, we quantified three interlocking constraints on cervical cancer control over the next 75 years: (i) most of the near-term cancer burden falls on women born before the country’s 2023 adolescent vaccination program could reach them, and can only be addressed through screening; (ii) opportunistic and even scaled screening in Nigeria is likely to be strongly mediated by education, meaning that the same women who missed vaccination are also less likely to reach screening, compounding the inequity; and (iii) infant vaccination could in principle offer a route around this cascade of correlated disadvantage, but only if it can achieve coverage and durability that meet or exceed the requirements set out in Equation 1, and only if routine infant delivery turns out to be more equitable in practice than school-based delivery. The findings on constraints (i) and (ii) rest on parameters calibrated to Nigerian data and vaccine efficacy estimates drawn from published trials in comparable settings, and we treat them as robust bases for near-term programme recommendations. The findings on constraint (iii), by contrast, are exploratory and hypothesis-generating: they are conditional on assumptions about infant HPV vaccine immunogenicity, durability across the ∼15–25-year interval to peak HPV exposure, and coverage achievable through Nigeria’s routine platform that current evidence cannot verify — no HPV vaccine trial has enrolled anyone below age 10, and no follow-up extends past approximately 11 years — and should be read accordingly throughout the discussion that follows.

The 70% screening coverage scenario we use as our comparator represents the WHO’s elimination target (WHO, 2020), not a forecast of what Nigeria can achieve in the near term. Current opportunistic screening in Nigeria reaches only ∼10–15% of women in their lifetime (16), so realizing the 70% target would require a roughly four-fold scale-up and substantial investment in cervical cancer screening infrastructure — trained providers, HPV DNA testing capacity, referral pathways for treatment, and demand generation. Population-level scale-up of screen-and-treat programmes has been achieved in comparable sub-Saharan African settings such as Zambia (28), demonstrating feasibility in principle, but the gap between what has been shown to be technically possible in demonstration programmes and what has been programmatically sustained at national scale remains large. Nigeria-specific analyses of cervical screening barriers (29) identify infrastructure constraints, geographic heterogeneity in access, and knowledge/awareness gaps as the dominant limiting factors. Results reported under the 70% scenario should therefore be read as the upper bound on what screening scale-up can contribute over the study horizon.

The coverage levels that an infant HPV vaccine program could attain would depend heavily on where in the routine schedule an infant HPV dose could be slotted: Nigeria’s own EPI schedule shows coverage falling sharply between earlier and later contacts, from 74% for BCG given at birth to just 36% for the second dose of measles-containing vaccine at 15 months (30), while DTP coverage was approximately 62– 63% through 2023–2025 (31). A dose timed early in infancy would likely reach higher coverage levels than one timed later, and if the delivery regimen required additional doses coverage would likely drop further. Assuming 62% coverage, our results show that infant delivery would avert 15–35% of cancers in vaccine-targetable cohorts, depending on effective VE at exposure (Figure 5), comparable to or worse than the reduction of around one-third averted under continued adolescent delivery at Nigeria’s current 60% aggregate coverage combined with WHO screening scale-up. Reaching 90% coverage for infant delivery may well require the same order of programmatic investment as reaching the 90% coverage target for adolescent delivery.

The routine immunization platform we would rely on for infant HPV delivery has its own substantial, well-documented coverage gaps linked to education and wealth: 2018 Nigeria DHS data show basic vaccination coverage (BCG, DTP-HepB-Hib, polio, and measles combined) ranging from 15% among children of mothers with no education to 47% among those with secondary education or higher (32), and Gavi’s 2023 Nigeria zero-dose analysis reports DTP3 coverage gaps exceeding 40 percentage points between the poorest and richest wealth quintiles (30). We are not aware of any study directly comparing HPV vaccine coverage equity to routine EPI coverage equity in the same population, so the claim that shifting delivery to infancy would reduce the education correlation we model for adolescent vaccination is, at present, an assumption rather than a demonstrated result, and the closest available Nigerian evidence suggests routine delivery is not, in practice, education-neutral. We therefore treat this as an open empirical question rather than a settled advantage of infant delivery. Delivery through routine childhood immunization may still offer advantages over school-based delivery, for instance, in reaching children before school-enrolment decisions are made, but reliable, equitable delivery is a hard problem for both strategies, not one that infant timing resolves by itself. Against whatever equity advantage infant delivery does offer must be set the cost of vaccinating two cohorts in parallel during any transition period, the possible need for multi-dose schedules if single-dose immunogenicity is insufficient at infant ages, and the opportunity cost of not investing in an adolescent-facing health platform that could deliver other adolescent services alongside HPV vaccination.

A core biological uncertainty for infant HPV delivery is whether protection persists across the ∼15–25-year interval between infant vaccination and peak HPV exposure. Numerous studies have now demonstrated HPV vaccine durability. Three (8–10) following the same Costa Rica Vaccine Trial cohort, vaccinated at ages 18–25 and followed for a median of ∼11 years: one (8) reports HPV16/18 infection durability, a second (9) reports cross-protection against non-vaccine types (HPV31/33/45) rather than vaccine-type durability, and the third (10) reports HPV16/18-associated CIN2+/CIN3 (precancer) for the trial’s three-dose recipients. Results from India (11) follow girls aged 10–18 for 9 years against a persistent-infection endpoint. The Kenyan single-dose trial (12) enrolled young women aged 15–20 and reported its primary persistent-infection endpoint at month 18. In short, no cited study measures durability against invasive cancer, none extends past ∼11 years, and none enrolled anyone younger than 10 (let alone infants). A related uncertainty is whether the efficacy evidence above, drawn entirely from participants aged 10 and older, applies at all to infants vaccinated in the first months of life. Similarly, the sufficiency of a single dose has been demonstrated in adolescent and young-adult recipients (11,12,33) but has not been tested in infants; a multi-dose schedule would raise per-fully-immunized-child costs and could reduce attainable coverage. These are precisely the empirical questions that our analysis is intended to motivate, not to resolve.

Our equity analysis reduces a multivariate gradient in vaccine and screening access to a single education-based odds ratio. Wealth, geography (particularly Nigeria’s north-south gradient in health service infrastructure), and healthcare access are all important determinants of HPV vaccine uptake and cervical screening access, and we did not model them as independent factors — quantitative estimates linking these determinants to HPV programme outcomes at the level of precision needed for the model were not available for Nigeria. Critically, these determinants are not independent of education in Nigeria; they are strongly co-linear predictors of health service use (34), so the joint gradient along any composite index of disadvantage would very likely be steeper than the education-only gradient we modelled. This means our equity finding — that residual cervical cancer burden concentrates on educationally-disadvantaged birth cohorts and that infant delivery does not automatically resolve this — should be read as a lower bound on the true equity concern, not an upper bound: a model with the true joint gradient would produce a larger residual burden and a larger equity gap, not a smaller one. Specific residual gradients that are unmodelled but likely amplifiers of the effect we do model include the north-south gradient in cervical screening infrastructure and the wealth-quintile gradient in DTP3 coverage that would carry over to any routine infant HPV dose (refs 30 and 32).

Our findings depend on several other simplifying assumptions. First, the vaccination-education coverage gap is represented by a single odds ratio, informed by regional campaign data rather than a Nigeria-specific or directly-fitted quantity; a smaller assumed gap would reduce the equity contrast between adolescent and infant delivery. We modeled a single-dose quadrivalent vaccine on the assumption that this remains the product supplied through Gavi; a switch to the 9-valent product would increase cross-protection and modestly widen the gap between vaccinated and unvaccinated cohorts. We did not have Nigeria-specific data on HPV genotype distribution in low-grade precancerous lesions, so our persistence estimates rely on genotype distribution in high-grade lesions. Similarly, our sexual behavior inputs draw on publicly-available survey data (35) that is subject to reporting bias and gaps.

Our results indicate that Nigeria’s introduction of HPV vaccination will halve the cervical cancer burden over the next 75 years, but that substantial residual burden will remain. For vaccine-targetable birth cohorts, infant delivery may be motivated by an intuitive equity argument, resting on the idea that routine childhood immunization would decouple HPV protection from school attendance and the educational determinants that also shape screening access. Our results suggest caution in applying this argument, especially given that the routine immunization platform also carries substantial coverage gaps linked to education and wealth. Whether infant delivery is nonetheless strategic hinges on empirical questions that only prospective studies and real programmatic data can resolve: how durable is single-dose infant immunity across two-plus decades, what fraction of infant HPV coverage Nigeria could sustain through the routine platform (and at what point in the schedule), how the resulting coverage gap compares to the one seen in school-based delivery, and how the delivery costs compare over a phased transition. Because these prerequisites are unresolved, our analysis does not support any policy recommendation contingent on infant HPV delivery at this time; we intend its findings on that strategy as hypotheses for the empirical work above to test, not as a basis for near-term programmatic decisions. The actionable near-term recommendation — which rests on parameters with strong empirical support — is sustaining and equitably scaling the adolescent vaccination program, with focused attention on out-of-school girls, together with a real investment in population-based screening for the pre-vaccine cohort.

## Data Availability

All data produced are available online at https://github.com/hpvsim/hpvsim_pxv_younger

## Declarations

## Ethics approval and consent to participate

Not applicable

## Consent for publication

Not applicable

## Availability of data and materials

The data and simulation code supporting the conclusions of this article is available in the https://github.com/amath-idm/hpvsim_pxv_younger repository.

## Clinical trial number

Not applicable

## Authors’ contributions

Study conception and design: JAC, CG. Model development: RMS, JAC. Writing the manuscript: RMS, UIB, JAC. All authors approved the final version.

## Acknowledgements

We thank Iacopo Baussano, Mark Jit, and Marc Brisson for commenting on an early version on this work.

